# Training-free generation of 4D CT and other rare acquisitions improves segmentation across modalities

**DOI:** 10.64898/2026.10.22.26365566

**Authors:** Christoph Sadée, Alex Dils, Navjeet Singh Hundal, Kyle Wan, Audrey Chun, Abhay Shukla, Ianto Lin Xi, Max Van Puyvelde, Katherine Hartmann, Preya Shah, Marc Hirschvogel, Wenhui Zhou, Cally Lin, Vineeth Gangaram, Ahmet Gorkem Er, Michał Seweryn, Wim Van Criekinge, Tarak Nath Nandi, Joel Ifenna Aniegbe, Luyao Shen, Adaobi Chikasi Ilo, Felix Immel-Sadée, Amaka Obiageli Nnamani, David L. Levin, Meng Hao, Elizabeth S McDonald, Nkechinyere Carina Achilefu, Nneka Ifeyinwa Iloanusi, Steven Paul Poplack, Brittany Z. Dashevsky, Scott M. Damrauer, Ifeoma Okoye, Olivier Gevaert

## Abstract

**Rationale:** Routine clinical imaging captures anatomy under a limited set of physiologic and acquisition conditions, leaving segmentation models with sparse examples of how lesions and surrounding tissues deform with compression, respiration, or vascular pulsation. Standard image augmentation expands training datasets but does not explicitly model the mechanics underlying these changes. Patient-specific biomechanical simulation offers an approach to generate anatomically consistent images and corresponding segmentation masks under conditions absent from the original scan, without additional imaging or training a generative model. We hypothesized that incorporating this mechanistically grounded variability into training improves segmentation across imaging modalities.

**Methods:** We evaluate across three segmentation tasks: breast cancer on 3D digital breast tomosynthesis, lung tumors on chest CT, and carotid plaque on ultrasound. From each routine scan, patient-specific biomechanical models generate acquisitions absent from the original study—a range of breast compressions, the respiratory cycle, and arterial wall distension across the cardiac cycle. The resulting displacement fields deform both images and masks. Segmentation models were trained with and without the generated data across a range of standard image augmentation techniques, evaluated via cross-validation and held-out test sets. Realism was assessed through blinded reader studies, and generated respiratory motion was validated against real 4D CT.

**Results:** Training on the generated acquisitions improved segmentation across modalities, increasing Dice scores by 6.6% for breast (p<0.05), 10.1% for lung (p<0.05), and 4.5% for carotid plaque (p>0.05) relative to the routine scans alone. Generated respiratory motion closely matched real gated 4D CT, with Dice of 0.94 ± 0.02 for lung volume overlap. Blinded readers discriminated generated from real images at near chance (AUC: breast 0.61, lung 0.41, carotid 0.46).

**Conclusions:** Mechanistic simulation generates imaging acquisitions that clinical systems do not supply, without training data or additional scanning, and improves AI-based disease detection across modalities.

## 1 Introduction

The development of accurate and generalizable medical AI requires large, heterogeneous training datasets to capture variation in disease phenotypes, yet such data are often scarce. In medical imaging, data availability is constrained by cost, privacy regulations, patient confidentiality and radiation exposure. Although AI-generated synthetic data has been proposed as a solution, it can introduce artifacts that degrade model performance, including hallucinations observed in both text- and image-based generative models [18, 38]. When exposed to data outside their training distribution, generative models may collapse or produce low-quality samples, particularly in underrepresented diseases or rare disease manifestations [20]. In addition, generative models can amplify biases present in the training data. Common evaluation metrics such as Fréchet Inception Distance (FID) and Kernel Inception Distance (KID) quantify distributional similarity between real and synthetic data [13, 26], but are insufficient in medical settings where anatomical and physiological plausibility are critical. More fundamentally, a generative model can only reproduce acquisitions represented in its training data. Acquisitions that clinical practice rarely or never performs such as dynamic expiratory CT (4D CT), largely confined to radiotherapy planning, or tomosynthesis across a range of compressions, which no clinical system acquires, therefore remain beyond reach.

As an alternative, we propose knowledge-based synthetic data generation, leveraging mechanistic models of human physiology to generate such acquisitions from a single routine scan, without any training data. Mechanistic knowledge can take many forms, including biological principles, mathematical models of tissue mechanics, organ motion, or treatment response [28, 29, 35]. Ordinary and partial differential equation (ODE/PDE) frameworks are widely available and used to model such processes and provide a principled means of constraining synthetic variation. Because distributional similarity metrics alone incompletely capture the relevance of synthetic data in biomedical applications, we evaluate realism through blinded clinician reader studies and assess downstream utility via segmentation performance.

We study three detection and segmentation tasks across distinct imaging modalities: breast cancer in 3D mammography, lung cancer in chest computed tomography (CT), and calcified carotid plaque in ultrasound. For each modality, we construct patient-specific physics-based models that simulate the underlying tissue deformation, generating acquisitions the original study did not capture. Finally, we validate realism with an expert readers study and in the instance of 4D CT, by comparing generated respiratory motion to real 4D CT.

## 2 Background and Methods

### 2.1 Scenario 1: 3D mammography and variation in breast compression

Breast cancer is the most common non-skin cancer among women [31]. In the US, screening and diagnosis are primarily performed using digital breast tomosynthesis (DBT), where the breast is compressed between a paddle and detector (below referred to as plates) while low-dose exposures are acquired over an x-ray tube arc and reconstructed into ∼1 mm slices (Figure 1A). Compression varies across patients and exams and contributes to study-to-study image variation, but it also reduces tissue overlap and improves image clarity and resolution, directly affecting lesion visualization. Here, we simulate clinically realistic variation in breast compression and the resulting change in image appearance to improve AI-based breast cancer detection and segmentation. An animation (breast.gif) of the simulated breast compressions is shown in the supplemental material.

**Figure 1.**
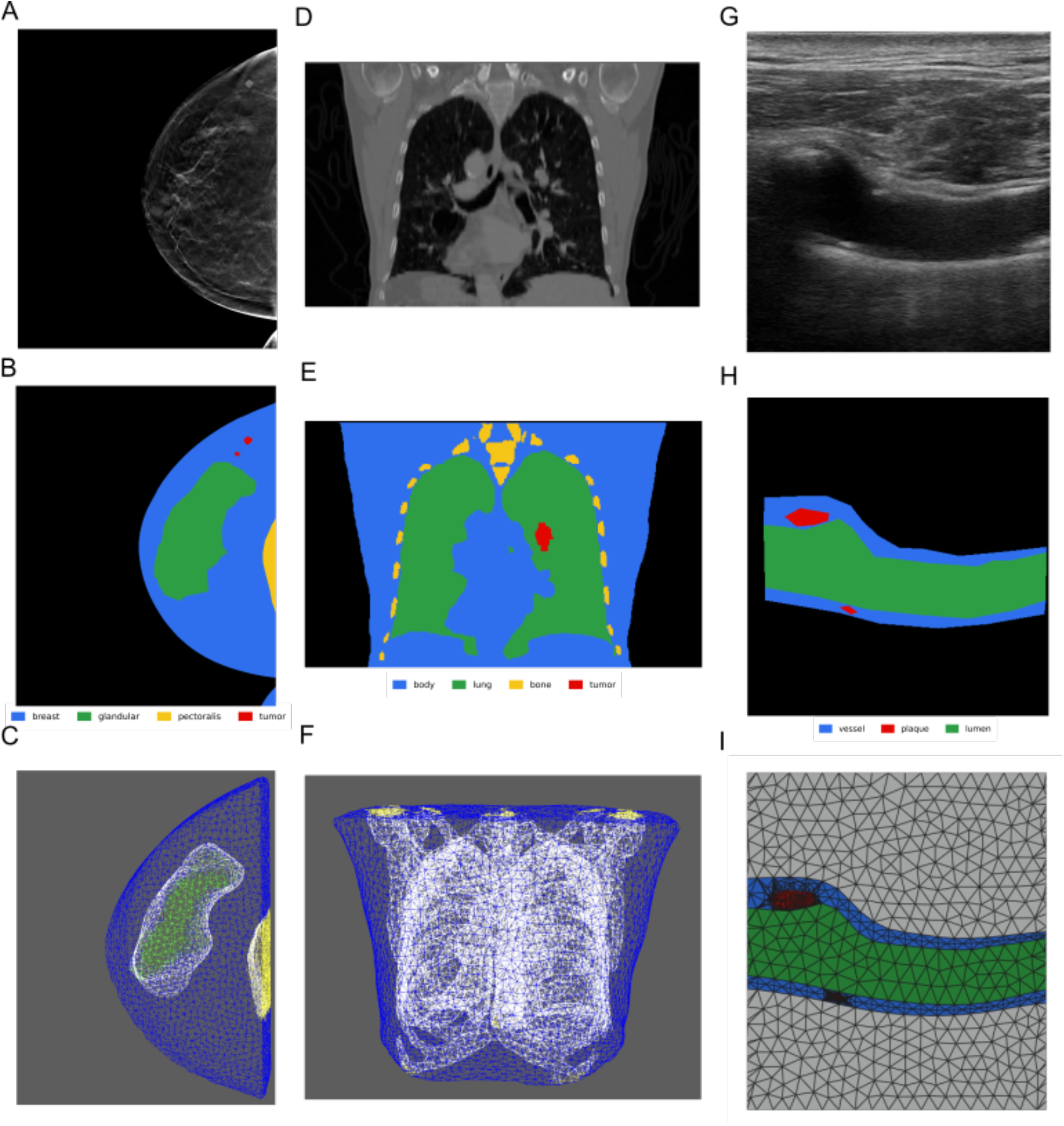
Representative images, manual segmentations, and corresponding 3D meshes across modalities. (A) 2D mammogram (MLO view) and (B) tissue segmentation (glandular: green; fat: red; pectoralis: yellow) with (C) derived 3D mesh. (D) Coronal chest CT with (E) lung, ribs, body wall, and tumor segmentation and (F) corresponding mesh. (G) Carotid ultrasound with (H) lumen and vessel wall segmentation outlines and (I) corresponding mesh including plaque.

#### 2.1.1 Dataset and Gold Standard Segmentation

We used the BCS-DBT dataset [6, 7], a publicly available digital breast tomosynthesis collection hosted on TCIA. From the full dataset (22,032 volumes from 5,060 patients), we selected biopsy-proven malignant cases, yielding 222 images from 158 patients (Table 4). Bounding box annotations provided with the dataset were used to guide manual lesion segmentation. We generated the first gold-standard tomosynthesis segmentation dataset by producing physician-reviewed manual lesion segmentations together with tissue segmentations of glandular tissue, fat, and pectoralis muscle (Figure 1B). Tissue labels were created by manually annotating a subset of images, training a 2.5D DeepLabV3 model to segment the remaining cases and subsequent manual corrections using ITK-SNAP [42].

#### 2.1.2 Physics model of breast compression

To simulate different compressions of the breast (breast.gif in supplemental material), we extracted the full 3D breast volume using the above-generated masks and constructed a tetrahedral finite-element mesh with subdomains corresponding to fat, glandular tissue, pectoralis muscle and lesion alongside their bounding surfaces. We also included an additional surface subdomain where the image truncates the body on the side of the breast. We then modeled compression as a quasi-static large-deformation hyperelasticity problem in which the unknown is the displacement field *u*: Ω → ℝ^3^ and the deformation map is *x* = *X* + *u*(*X*):

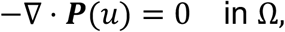

We used a compressible Neo-Hookean constitutive law. Let *F* = *I* + ∇*u* be the deformation gradient, *J* = det(*F*), and *I*_*c*_ = tr(*F*^⊤^*F*). The strain-energy density is:

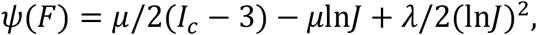

and the first Piola–Kirchhoff stress is ***P*** = ∂ψ/ ∂*F*. The Lamé parameters (λ, *µ*) were assigned piecewise-constantly by tissue type using standard conversions from (*E, ν*) with tissue properties as defined in supplemental Table 5.

To model contact between deformable breast tissue and the rigid compression plates, we used a penalty regularization approach [25]. We represented the plates as two rigid geometries (top and bottom) that move symmetrically along the compression axis.

When the deformed surface penetrates a plate, a penalty traction is applied along the plate normal:

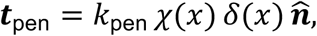

where *x* = *X* + *u* is the deformed position, *χ*(*x*) is an indicator active only when penetration occurs, *δ*(*x*) is the signed penetration depth, and 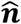 is the plate normal. Following our implementation, the penalty stiffness was scaled with mesh resolution via *k*_pen_ = *C*_pen_ *E*_eff_/*h*, where *h* is a characteristic element size and *E*_eff_ is an effective Young’s modulus derived from (λ, *µ*).

#### 2.1.3 Simulation and image generation

The simulation was implemented in FEniCSx [4] using a nonlinear finite element formulation of 3D hyperelasticity. We used continuous first-order (P1) Lagrange elements to discretize *u* on the 3D mesh (Figure 1C). Compression was applied by incrementally increasing the symmetric plate displacement over multiple load steps; at each step, the nonlinear variational problem was solved with Newton’s method using PETSc linear solvers, and contact forces were incorporated via the penalty traction described above.

The resulting displacement field *u*(*x*) was exported and used to warp the original tomosynthesis volume using scikit-image [36]. For each voxel position *x*, the warped location was computed as *x*^’^ = *x* + *u*(*x*), and the intensity at *x*^’^ was interpolated from the original image using linear interpolation. Areas outside the mesh were removed to prevent artifacts. This procedure yielded physically plausible synthetic views under increased compression (see supplemental breast.gif), which were used as synthetic data in the reader study and AI segmentation tasks.

### 2.2 Scenario 2: Lung CT with variable respiration

Lung cancer is the second most common cancer and the leading cause of cancer death in the United States [32]. Current clinical practice relies heavily on computed tomography (CT) of the chest, a 3D imaging modality capturing the lung and surrounding tissue (Figure 1D) for detection, diagnosis and monitoring of lung lesions.

During imaging, patients are asked to hold their breath resulting in capture of lung volumes at inspiration. Currently, lesions are almost exclusively identified and measured manually, which is time-consuming and labor-intensive. Accordingly, there are numerous efforts for automated segmentation methods to aid medical professionals in identifying and measuring lesions [40]. However, limited labeled data and high appearance variability remain key challenges. Here, we simulate clinically realistic variation in lung inspiration and the resulting change in image appearance to improve AI-based lung cancer detection and segmentation.

#### 2.2.1 Dataset and Segmentations

We used the LOTUS chest CT dataset, derived from NSCLC-Radiomics with additional annotations from the MAASTRO clinic [1]. The dataset contains one CT volume per patient (300 publicly available) with expert-delineated gross tumor volume and body contours (Figure 1E). Lung and bone segmentations were obtained using TotalSegmentator [39], selecting labels corresponding to lung parenchyma and bony structures. Provided tumor and body contours were converted into voxel-wise segmentations using standard rasterization tools [5, 21]. To ensure a watertight simulation domain required by the physics model, internal cavities were filled and all classes were merged into a single composite segmentation.

#### 2.2.2 Physics Model of Respiratory Motion

Respiratory motion was simulated using a patient-specific 3D finite-element model with subdomains for body, ribs, lung, and tumor. Breathing was modeled as a quasi-static large-deformation hyperelasticity problem, solving for the displacement field *u*: Ω → ℝ^3^ under mechanical equilibrium,

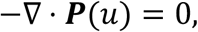

where ***P*** is the first Piola–Kirchhoff stress. Tissue mechanics were described using a compressible Neo-Hookean constitutive law, with material parameters assigned per tissue class from (*E, ν*) (Table 5). Respiratory motion was driven by a sinusoidal pressure traction applied to the lung boundary, *p*(*t*) = *p*_0_sin(2*πft*) (default *p*_0_ = −2400 Pa, *f* = 18/60 Hz), evaluated over 24 time steps per breathing cycle. Ribs were treated as rigid by imposing zero-displacement boundary conditions.

#### 2.2.3 Simulation and image generation

The simulation was implemented in FEniCSx using a nonlinear finite element formulation of 3D hyperelasticity on the patient-specific mesh (Figure 1F). The sinusoidal pressure load was advanced over 24 time steps, and at each step the nonlinear variational problem was solved using Newton’s method to obtain the displacement field *u*(*X, t*). The resulting displacement field was exported (with time indexing) and used to warp the original CT volume and all corresponding segmentation masks, yielding label-consistent augmented data.

### 2.3 Scenario 3: Ultrasound and arterial wall distension of the carotid

Carotid ultrasound is a widely used, noninvasive modality for assessing atherosclerotic plaque burden and estimating stroke risk by measuring luminal (vessel) narrowing in the common and internal carotid arteries. During the cardiac cycle, pulsatile blood flow induces cyclic arterial wall motion that alters vessel geometry, lumen diameter, and plaque appearance; a still B-mode image captures the vessel at a single, arbitrary point in that cycle. Here, we simulate clinically realistic variation in arterial wall distension and the resulting change in image appearance to improve AI-based plaque detection and segmentation.

#### 2.3.1 Dataset and Gold standard Segmentations

The Carotid Ultrasound Boundary Study (CUBS) dataset [23], accessible via kaggle, is a collection of 500 B-mode ultrasound images of the common carotid artery (Figure 1G). We excluded 100 synthetically generated images, due to unrealistic appearances. A radiologist manually segmented each image [37] to denote the lumen, vessel wall, and plaque if present as shown in Figure 1H.

#### 2.3.2 Fluid–Structure Interaction (FSI) Model

An extended description is provided in Supplementary Section alongside boundary conditions and numerical implementation 6.1.3. We model carotid wall distension and plaque motion using a standard fluid–structure interaction (FSI) formulation that couples a hyperelastic vessel wall to incompressible blood flow in an Arbitrary Lagrangian– Eulerian (ALE) frame [10, 11]. The vessel wall displacement ***u*** on the reference solid domain 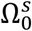 satisfies finite-strain elastodynamics,

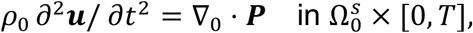

where ***P*** is the first Piola–Kirchhoff stress derived from a hyperelastic strain-energy density. Blood flow in the deforming fluid domain Ω^*f*^ is modeled as incompressible Newtonian flow,

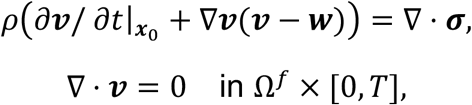

with Cauchy stress ***σ*** = −*p****I*** + *µ*(∇***v*** + ∇***v***^T^). At the fluid–solid interface Σ, kinematic continuity and traction balance are enforced,

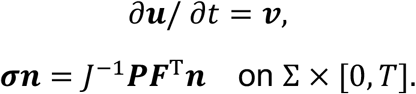

All simulations were computed using Ambit on the FEniCSx framework with PETSc-based nonlinear and linear solvers [2, 3, 14].

### 2.4 Training AI segmentation models

To evaluate the impact of physics-based synthetic data, we trained segmentation models with and without it across all modalities. For lung CT, two synthetic acquisitions were generated by increasing peak exhalation pressure; for breast tomosynthesis, four compression levels were simulated by decreasing plate distance (2.5, 5, 7.5, and 10 mm); and for carotid ultrasound, three cardiac-phase samples were selected at peak systole, diastole, and the dicrotic notch, together with an additional constant-flow profile of 85 cm/s. Generated 3D volumes were post-processed with modality-specific windowing, and 2D slices containing the largest lesion extent were extracted together with the corresponding segmentation masks.

Data were split into five cross-validation folds with an additional held-out test set per dataset, ensuring that all synthetic images remained within the same fold as their source image to prevent leakage. Synthetic images were excluded from test sets and from baseline training runs. Images were resized to fixed input resolutions while preserving aspect ratios (breast: 650 × 800, lung: 500 × 400, carotid: 800 × 600).

All models were trained for 200 epochs using a batch size of 32 and the Adam optimizer with a combined cross-entropy and Dice loss. A base learning rate of 10^−4^ with 5-epoch linear warm-up and cosine decay to 10^−6.^ was used; class imbalance was addressed via dataset-derived log-scaled class weights. For each fold, the best checkpoint was selected based on validation performance and evaluated on the test set, yielding five scores per experiment.

For lung CT, we additionally evaluated generalization across architectures, including DeepLabV3 with EfficientNet-B4, ResNet-50, and ConvNeXt backbones, as well as SegFormer. The effect of standard image-space transformations was assessed both independently and in combination with physics-based augmentation. All experiments were performed on NVIDIA RTX 6000 Pro GPUs.

## 3 Results

### 3.1 Generation of acquisitions absent from the original study

From a single routine scan, the framework produces acquisitions absent in the originating study. For lung CT, one breath-hold volume yields a complete respiratory cycle sampled at 24 phases per patient, finer than the ten phases of the gated 4D CT cohort used for validation and obtained without the additional exposure a gated acquisition requires. For breast tomosynthesis, one compressed volume yields the breast across a range of plate separations (2.5–10 mm); because compression is advanced incrementally over load steps, any intermediate state can be recovered, of which four were sampled for training. For carotid ultrasound, a single frame yields the vessel across the cardiac cycle. In each case the acquisition is derived from data already collected in routine care, requiring no repeat imaging and no additional radiation.

The two radiographic acquisitions differ in whether a reference standard exists. Respiratory motion can be compared directly against gated 4D CT, which is acquired clinically, if rarely, and we report that comparison in Section 3.4. Tomosynthesis at multiple compressions has no such counterpart: no clinical system images a patient at more than one compression, because each state would require its own exposure. For that acquisition no reference data exist against which to validate, and perceptual judgment by expert readers is the available standard. Generation is inexpensive in either case, requiring ∼8 min per 3D volume on CPU and no specialized hardware (Section 6.2.5).

### 3.2 Anatomical plausibility of generated images

We assess the realism of the proposed physics-based synthetic data through qualitative visualization using animations (breast.gif and lung.gif in supplemental material) and deformed images.

Figure 2 presents examples of physics-based synthetic data for breast tomosynthesis, lung CT, and carotid ultrasound. Across all modalities, the induced deformations are spatially coherent and constrained by the underlying physiological process modeled. In the breast scenario, simulated increases in compression produce heterogeneous tissue expansion and lesion displacement while preserving structural constraints, including limited motion of the stiffer pectoralis muscle in contrast to the more compliant fatty and glandular tissue. In the lung scenario, respiratory motion induces lesion displacement within the lung volume together with physiologically plausible downward motion of the diaphragm, while surrounding bones and body wall remain comparatively stationary. In the carotid artery scenario, the cardiac cycle produces outward movement of the arterial wall, together with localized plaque displacement at peak systole.

**Figure 2.**
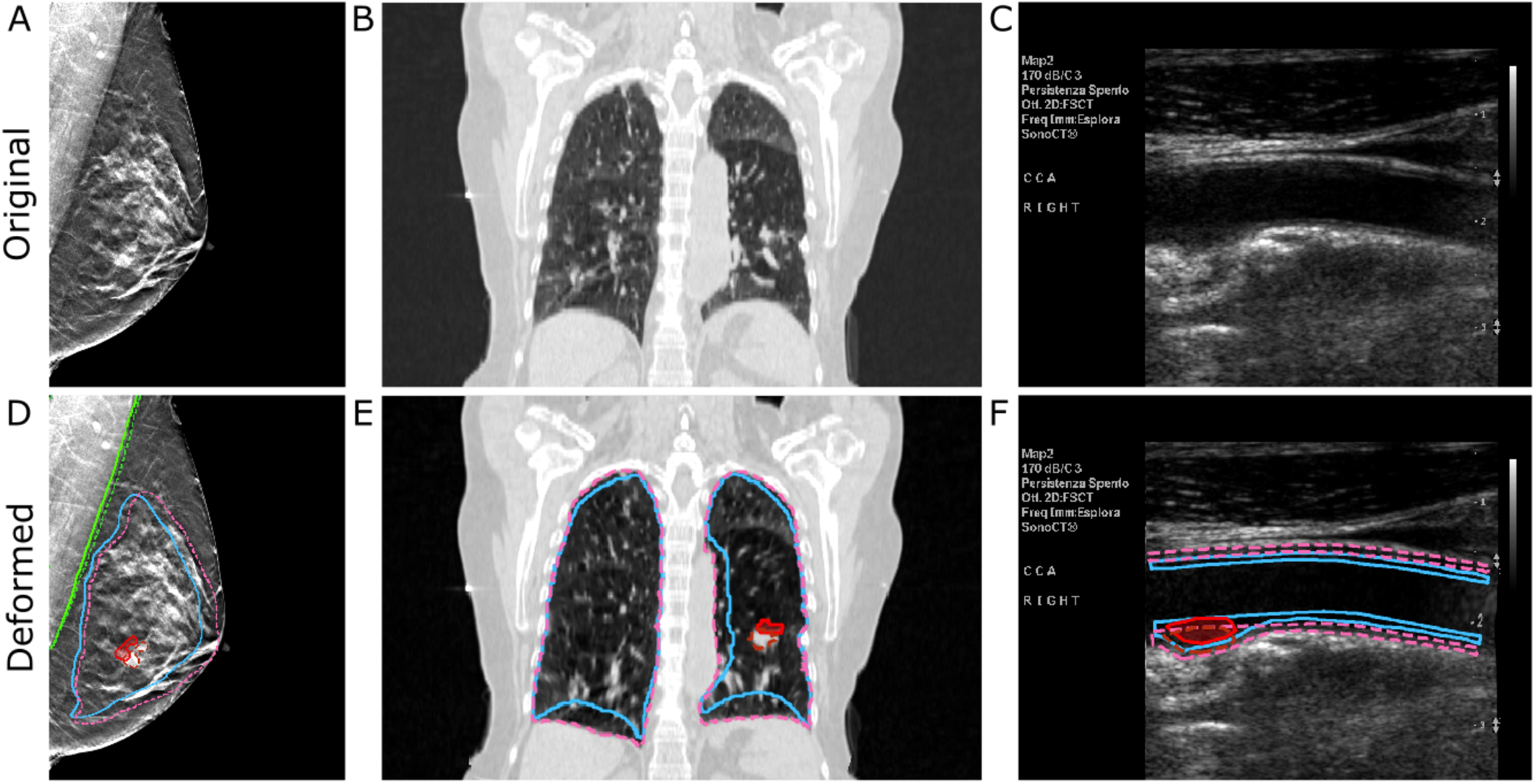
Physics-based synthetic data for breast DBT, lung CT, and carotid ultrasound. (A,B,C) Original images (D,E,F) Corresponding generated images; solid/dashed outlines denote pre-/post-deformation.

Across all cases, the generated deformations selectively modify target anatomical structures while preserving global anatomical consistency, providing a qualitative demonstration that the physics-based models generate realistic, label-consistent deformations suitable for data augmentation. This contrasts with other augmentation strategies, which may introduce arbitrary warping (see Figure 4). Because physics-based deformations arise from models that closely mimic the underlying biology, they preserve anatomical relationships while introducing physiologically meaningful variation.

### 3.3 Reader Study

To assess the perceptual realism of the proposed physics-based synthetic data, we conducted a blinded reader study in which expert physicians were asked to classify images as real or synthetic. Readers routinely interpret the corresponding imaging modalities as part of their clinical workflow. Performance was summarized using sensitivity, specificity, and estimated AUC, noting that these metrics reflect qualitative perceptual judgments rather than objective diagnostic ground truth. For breast DBT, synthetic images with visible edge artifacts at the chest boundary were excluded; for carotid ultrasound, a warped on-image scale bar was cropped out before review.

Across all modalities, reader performance was near chance. Synthetic images were correctly identified in 60% of mammograms, 52% of chest CT images, and 42% of carotid ultrasound images (Table 1). Corresponding sensitivity and specificity values were balanced and low across modalities, and inter-reader agreement was poor to slight, with Fleiss’ *κ* values close to zero (Table 6). Together, these results indicate that expert readers were generally unable to reliably distinguish physics-based synthetic images from real clinical images.

**Table 1.**
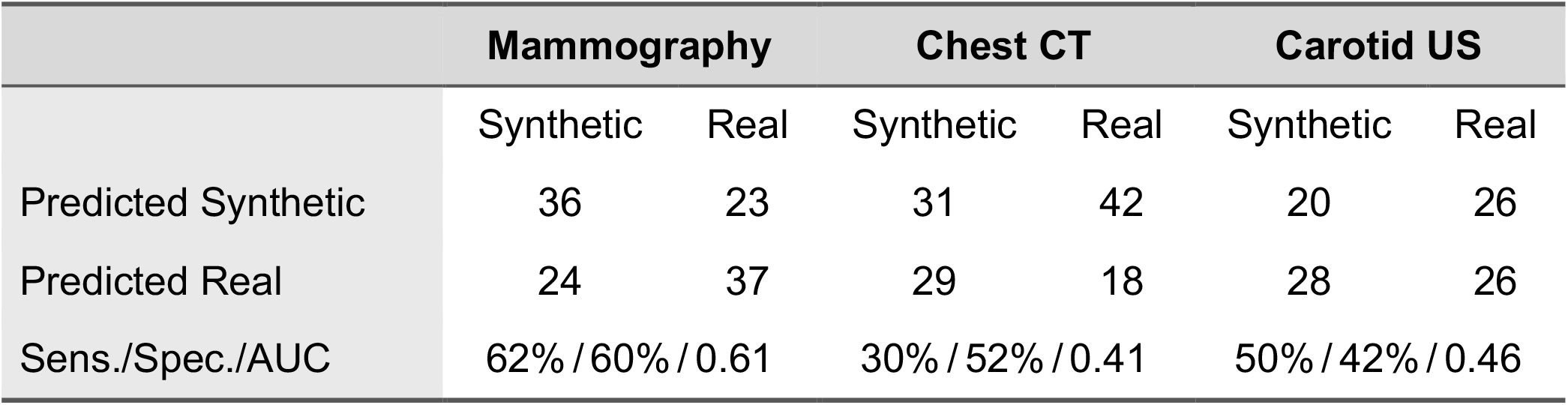
Confusion matrices for predicted versus actual labels (synthetic vs. real) across imaging modalities. Sensitivity, specificity, and estimated AUC for each modality are listed below.

Performance for lung and carotid was essentially at chance. Performance above chance for mammography (AUC = 0.61) was driven primarily by a single reader (Reviewer 3), who achieved an estimated AUC of 0.83 as seen in Table 7. Upon inquiry, this reader identified a localized artifact at the image border (20 × 20 pixels) of full-resolution mammograms (1889 × 2457 pixels) that did not affect the realism of the overall breast tissue or areas used for diagnostic purposes. When excluded, mammography performance returned to near chance (estimated AUC = 0.53). Overall, the combination of near-chance classification performance and low inter-reader agreement supports the conclusion that physics-based synthetic images are perceptually indistinguishable from real images for expert readers.

### 3.4 Validation of Simulated Respiratory Motion

We next evaluate the ability of the proposed simulation to capture physiologically realistic motion, using lung respiration as a case example due to the availability of a separate 4D chest CT cohort.

Simulated lung motion was compared against ground-truth masks from gated 4D chest CT scans of twenty patients from TCIA, spanning ten increments of an individual breathing cycle (see lung.gif animation in supplemental material). Starting from the CT acquired at the initial respiratory phase, the simulation predicted the remaining nine increments, and the resulting lung masks were evaluated against ground truth. For each patient, intrathoracic pressure was optimized to match the observed lung-volume trajectory varying between 2.4–4.5 kPa. This flexibility indicates that the simulation can be readily adapted to generate both inter- and intra-patient variability, enabling multiple physiologically plausible augmentations rather than a single deterministic deformation, which can be leveraged for more robust AI training.

Across all patients, simulated lung-volume trajectories closely matched the ground truth, with Dice scores of at least 0.90 (Table 8). Spatial discrepancies were further quantified using the average symmetric surface distance (ASSD) and Hausdorff distance (HD).

Across all respiratory increments and patients, ASSD remained below 3 mm, with an average HD of 22.8 mm. The largest deviations occurred at peak expiration and are illustrated for Patient 1 in Figure 3. In these cases, false positives were primarily located along the outer lung boundary, while false negatives clustered in between the lung lobes -the mediastinum. These patterns are consistent with the model’s simplifying assumption of fully rigid bone structures, which reduces lung contraction especially at peak expiration. Despite this, mean surface error remained below 3 mm even in the least optimal case.

**Figure 3.**
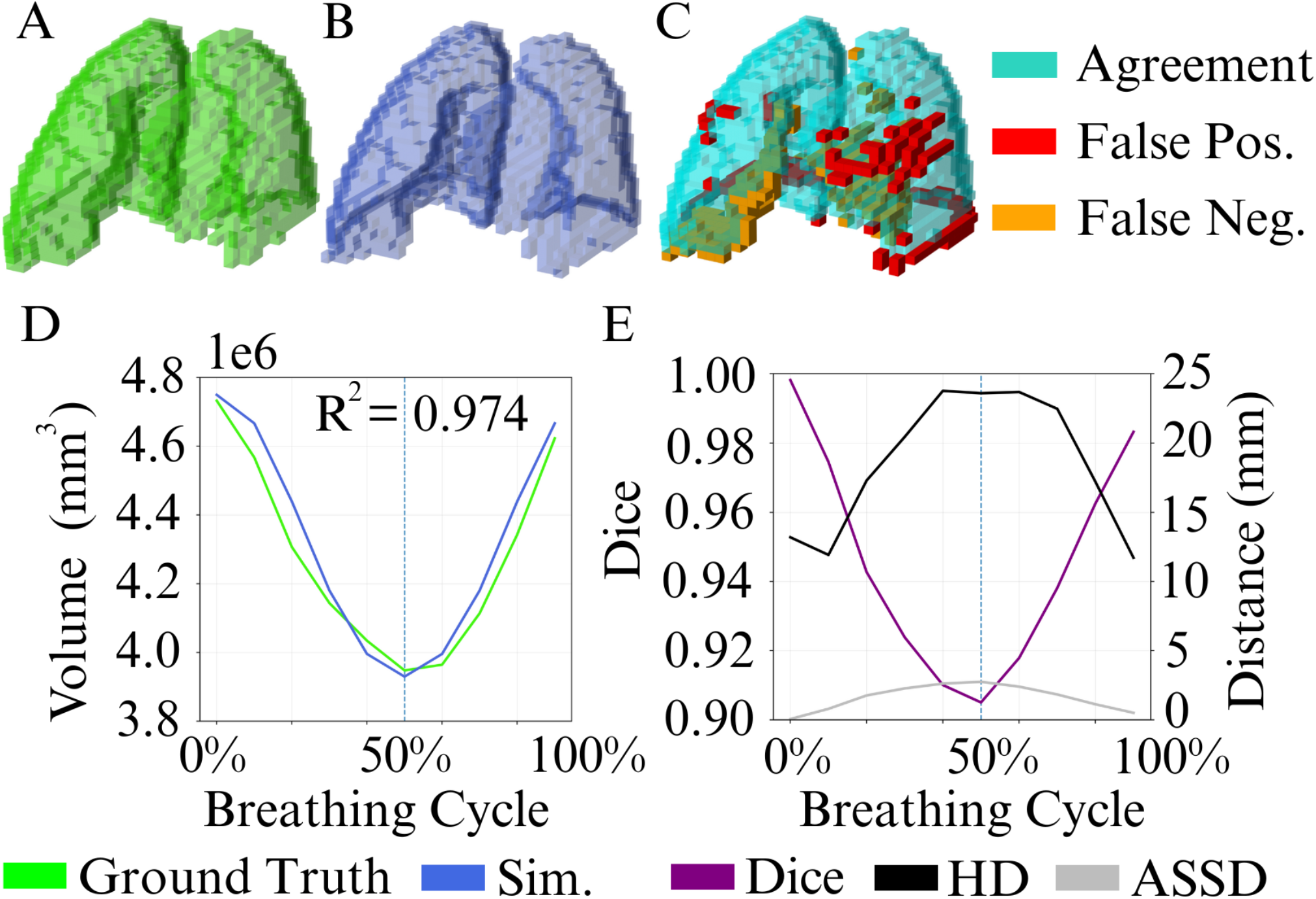
Qualitative and quantitative comparison between ground-truth and simulated lung masks across respiratory cycle for patient 1. Each 3D rendering is downsampled to a voxel size of 10 mm^3^ to highlight larger deviations. A) Ground truth 3D lung mask. B) Simulated 3D lung mask. C) Combined visualization highlighting overlap and disagreement. D) Ground-truth and simulated lung volume trajectories across respiratory cycle. E) Dice, HD, and ASSD metrics across respiratory cycle. The vertical line in D and E indicates the breathing stage captured in A, B and C.

**Figure 4.**
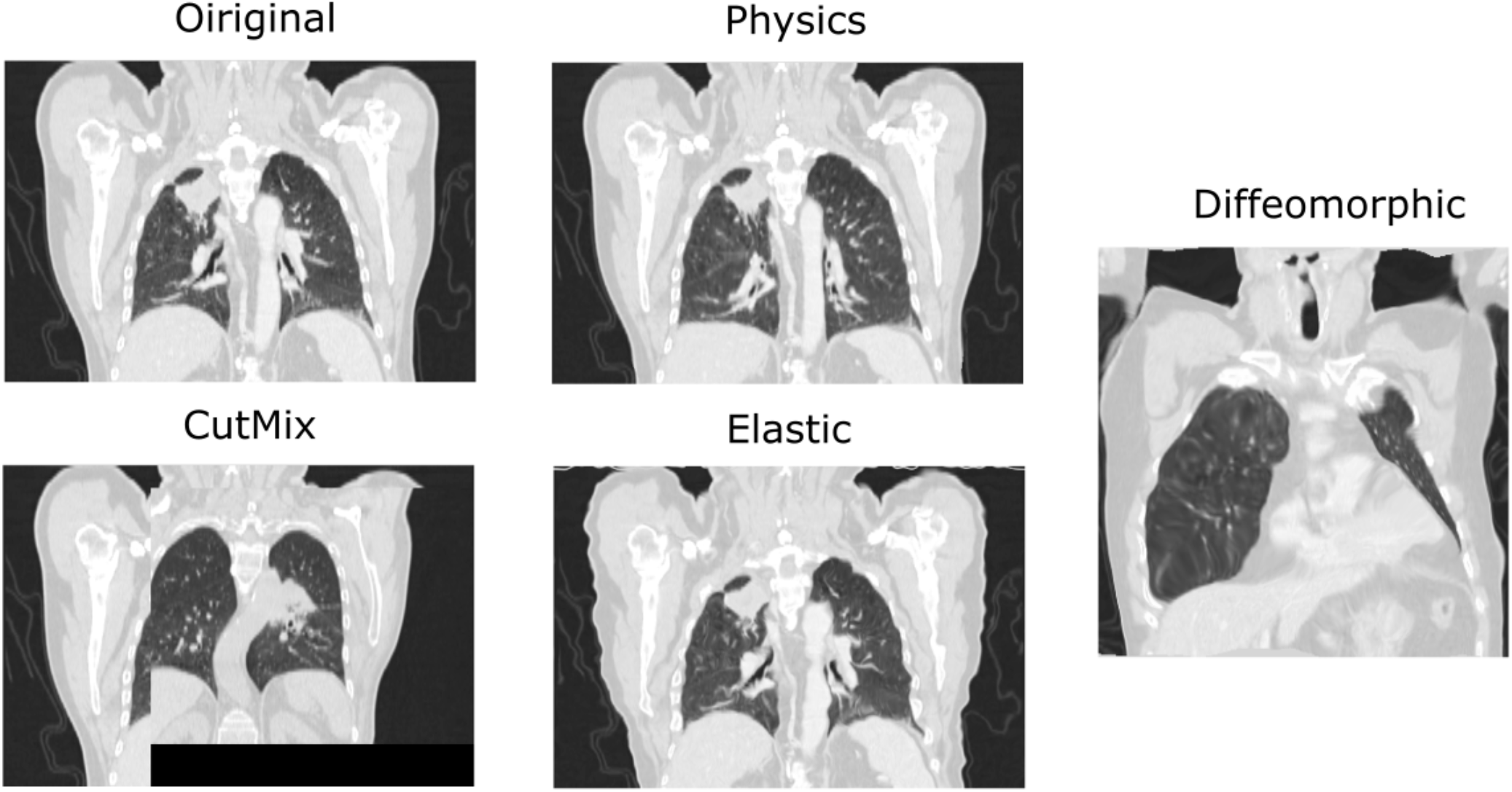
Shown are examples of physics-based augmentation compared to other augmentation strategies such as CutMix and Elastic deformation and Diffeomorphic registration. For diffeomorphic registration the original image is aligned to another image in the dataset and a deformation field is computed. This deformation field is then applied to the original image to generate the augmented image half way between the original and other image.

Although no direct benchmarks exist for this public dataset, our approach achieved competitive performance relative to prior work evaluated on other datasets, including the lowest mean Hausdorff distance (22.80 ± 8.44 mm) among the compared methods (Table 10). While deformable registration approaches [12, 27] achieve higher Dice scores and lower ASSD, they require access to full 4D CT sequences and therefore cannot generate synthetic motion from a scan acquired at a single time point. In contrast, our method relies on a single, clinically common CT acquisition, enabling scalable augmentation for downstream training. Taking into consideration differences in datasets and evaluation protocols, our approach achieves higher Dice scores than the physics-based lung biomechanical simulation of [16] (0.94 ± 0.02 vs. 0.85 ± 0.06).

Performance even remained high in an ablation study, simulating each patient without volume optimization (pressure fixed to 2.4 kPa) and no global alignment, see Table 9 (mean Dice: 0.94, ASSD: 1.58, HD: 22.32)

Overall, these results demonstrate that physics-based simulations can generate accurate and physiologically plausible respiratory motion from a single CT acquired at a single time point, supporting their use as a practical and scalable augmentation strategy.

### 3.5 AI Segmentation Improvements

We first evaluate physics-based augmentation against standard image-space transformations (color jitter, rotation, flipping) and advanced alternatives (elastic deformation, CutMix [41]) using a fixed EfficientNet-B4 DeepLabV3 architecture across breast, lung, and carotid datasets. Each augmentation is tested alone and in combination with physics-based augmentation (Table 2).

**Table 2.** Dice scores for lesion/plaque segmentation across modalities (EfficientNet-B4). Each pair of rows compares a baseline augmentation (top) against the same augmentation combined with physics-based augmentation (bottom, + P.). p-values are FDR-corrected.

|  | *Breast* | *p* | *Lung* | *p* | *Carotid* | *p* |
| --- | --- | --- | --- | --- | --- | --- |
| None | 0.623 | **.001** | 0.515 | **.001** | 0.306 | .947 |
| Physics | **0.689** |  | **0.616** |  | **0.351** |  |
| Jitter | 0.573 | **.001** | 0.540 | **.001** | 0.355 | .947 |
| Jitter + P. | **0.670** |  | **0.615** |  | **0.387** |  |
| Rotate | 0.667 | **.002** | 0.595 | $< 1e-3$ | 0.372 | .947 |
| Rotate + P. | **0.707** |  | **0.671** |  | **0.388** |  |
| Flip | 0.624 | **.004** | 0.555 | $< 1e-3$ | **0.354** | .947 |
| Flip + P. | **0.687** |  | **0.643** |  | 0.330 |  |
| Elas. Defo. | 0.638 | **.027** | 0.568 | $< 1e-3$ | 0.349 | .947 |
| Elas. Defo. + P. | **0.677** |  | **0.638** |  | **0.368** |  |
| Cutmix | 0.608 | .172 | 0.581 | $< 1e-3$ | 0.362 | .947 |
| Cutmix + P. | **0.645** |  | **0.628** |  | **0.366** |  |

Across all three datasets, physics-based augmentation consistently improves segmentation performance relative to no augmentation. Performance gains are most pronounced for breast tomosynthesis and lung CT, with Dice score improvements of 6.6% (p < 0.05) and up to 10.1% (p < 0.05), respectively. For carotid ultrasound, physics-based augmentation yields a consistent but non-significant improvement of 4.5% (p > 0.05), likely reflecting the limited number of plaque-positive samples (38 cases) available for synthetic data generation. Despite this, the overall trend mirrors that observed for breast and lung imaging.

Physics-based augmentation consistently outperforms standard image-space transformations when applied in isolation, while also complementing them when used in combination. For lung CT, physics-based augmentation alone (Dice 0.616) outperforms jitter (0.540), rotation (0.595), and flipping (0.555), a pattern that similarly holds for breast tomosynthesis, where physics-based augmentation alone exceeds all standard transformations. Notably, for both lung CT and breast tomosynthesis, the highest performance is achieved when combining physics-based augmentation with rotation (Dice 0.671 and 0.707, respectively), indicating that physics-based and standard augmentations provide complementary sources of variation rather than redundant transformations. For carotid ultrasound, although differences are not statistically significant, the combination of physics-based augmentation and rotation likewise yields the highest observed performance among the evaluated transformations, underscoring that physics-based augmentation can be effectively integrated with existing augmentation strategies to further improve model performance.

The pattern holds for more advanced image-space transformations. Elastic deformation applies a random smooth displacement field to the image, while CutMix replaces a random rectangular region with a patch from another training image [41]. For both breast and lung, neither elastic deformation nor CutMix outperforms physics-based augmentation alone. In all cases, combining these transformations with physics-based augmentation further improves performance.

Next, we assess whether performance gains from physics-based augmentation generalize across different backbones and architectures using lung CT as a trial case (Table 3). Physics-based augmentation improves Dice scores across all evaluated backbones (EfficientNet-B4, ResNet-50, ConvNeXt) as well as the SegFormer architecture. Peak performance (Dice 0.681) is achieved using ResNet-50 when combining physics-based augmentation with flipping, beating much newer architectures such as SegFormer. Across architectures, physics-based augmentation alone consistently outperforms standard augmentations, with the exception of SegFormer, where the largest gains arise from combining both. Advanced transformations occasionally match physics-based augmentation (e.g., ConvNeXt: CutMix 0.632 ≈ Physics 0.632), but combining them yields further improvement (CutMix + P. 0.644), reinforcing the complementarity of physics-based augmentation.

**Table 3.** Dice scores for the lung dataset across backbones and SegFormer. Each pair of rows compares a baseline augmentation (top) against the same augmentation combined with physics-based augmentation (bottom, + P.). p-values are FDR-corrected. See Section 6.2.4 for details.

| Augmentation | Eff. B4 | p | ResNet | p | ConvNeXt | p | SegFormer | p |
| --- | --- | --- | --- | --- | --- | --- | --- | --- |
| None | 0.515 | .001 | 0.605 | .018 | 0.593 | .001 | 0.593 | .001 |
| Physics | 0.616 |  | 0.658 |  | 0.632 |  | 0.635 |  |
| Jitter | 0.540 | .001 | 0.616 | .018 | 0.601 | .045 | 0.606 | .010 |
| Jitter + P. | 0.615 |  | 0.654 |  | 0.625 |  | 0.666 |  |
| Rotate | 0.595 | <1e-3 | 0.632 | .020 | 0.617 | .001 | 0.649 | .566 |
| Rotate + P. | 0.671 |  | 0.667 |  | 0.661 |  | 0.663 |  |
| Flip | 0.555 | <1e-3 | 0.641 | .001 | 0.612 | .002 | 0.646 | .137 |
| Flip + P. | 0.643 |  | 0.681 |  | 0.662 |  | 0.673 |  |
| Elastic def. | 0.568 | <1e-3 | 0.636 | .020 | 0.597 | .040 | 0.634 | .109 |
| Elastic def. + P. | 0.638 |  | 0.674 |  | 0.635 |  | 0.655 |  |
| CutMix | 0.581 | <1e-3 | 0.636 | .221 | 0.632 | .078 | 0.605 | .068 |
| CutMix + P. | 0.628 |  | 0.660 |  | 0.644 |  | 0.638 |  |

**Table 4.** Summary of datasets used for synthetic data generation, segmentation, and reader study. Tumor/plaque masks were manually annotated and reviewed by a radiologist. CT anatomical masks were generated with TotalSegmentator (others manually); the body class denotes all remaining anatomy. Only 38 images in the carotid dataset contained plaque.

|  | Breast | Lung | Carotid |
| --- | --- | --- | --- |
| **Dataset Name** | BCS-DBT | LOTUS | CUBS |
| **Source** | TCIA | LOTUS | kaggle |
| **Datatype** | 3D Mammo. | Chest CT | Ultrasound |
| **Sample Size** | 209 | 293 | 400 |
| **Segmentation** | fat<br>pectoralis<br>glandular<br>tumor | body<br>bone<br>lung<br>tumor | body<br>vessel wall<br>lumen<br>plaque |

**Table 5.** Tissue properties used for the physics-based simulations of breast compression and lung respiration. E is the Young’s modulus and ν the Poisson ratio.

| Scenario | Tissue | $E$ (kPa) | $\nu$ |
| --- | --- | --- | --- |
| Breast | Fat | 0.75 | 0.45 |
|  | Glandular | 2.25 | 0.45 |
|  | Pectoralis | 62.2 | 0.40 |
|  | Tumor | 3.75 | 0.45 |
| Lung | Body | 10.0 | 0.45 |
|  | Lung | 6.12 | 0.40 |
|  | Tumor | 12.73 | 0.40 |
| | Bone | $1.39 \times 10^7$ | 0.30 |

**Table 6.** Reviewer expertise and inter-rater agreement (Fleiss’ Kappa) across the three imaging modalities. Reviewer experience is reported in years (y). All reviewers are consultant radiologists, with the exception of ^*^ a consultant vascular surgeon.

|  | Mammography | Chest CT | Carotid US |
| --- | --- | --- | --- |
|  | 30 images | 30 images | 25 images |
| Reviewer 1 | >5y | <5y | >5y* |
| Reviewer 2 | >20y | <5y | >5y |
| Reviewer 3 | <5y | >20y | >5y |
| Reviewer 4 | >20y | <5y | >15y |
| Fleiss' Kappa | -0.03 (poor) | -0.06 (poor) | 0.03 (poor) |

**Table 7.**
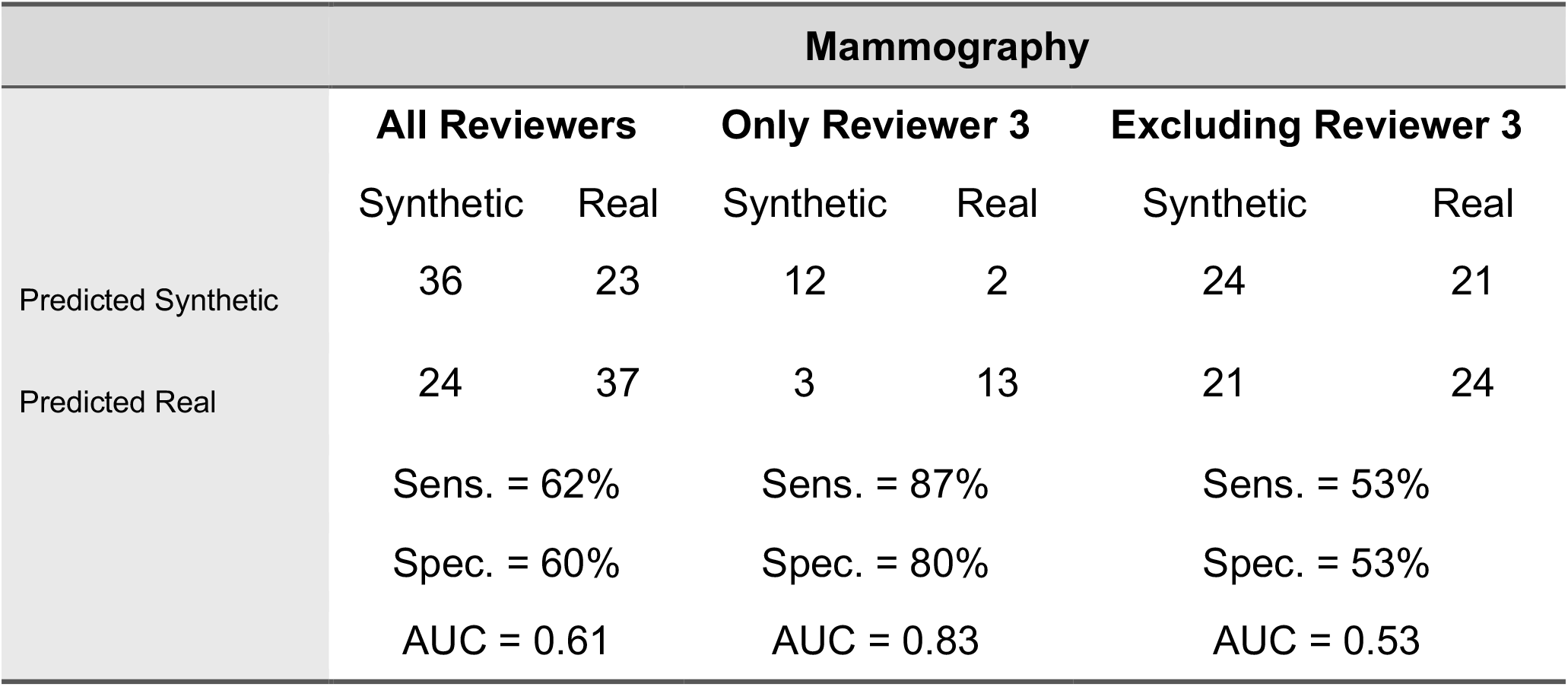
Confusion matrices for predicted versus actual labels (synthetic vs. real) for Mammographies. Results are shown for all reviewers combined, Reviewer 3 only, and all reviewers excluding Reviewer 3. Sensitivity, specificity, and estimated AUC for each category are listed below.

| Mammography |  |  |  |  |  |  |
| --- | --- | --- | --- | --- | --- | --- |
|  | All Reviewers |  | Only Reviewer 3 |  | Excluding Reviewer 3 |  |
|  | Synthetic | Real | Synthetic | Real | Synthetic | Real |
| Predicted Synthetic | 36 | 23 | 12 | 2 | 24 | 21 |
| Predicted Real | 24 | 37 | 3 | 13 | 21 | 24 |
|  | Sens. = 62% |  | Sens. = 87% |  | Sens. = 53% |  |
|  | Spec. = 60% |  | Spec. = 80% |  | Spec. = 53% |  |
|  | AUC = 0.61 |  | AUC = 0.83 |  | AUC = 0.53 |  |

**Table 8.**
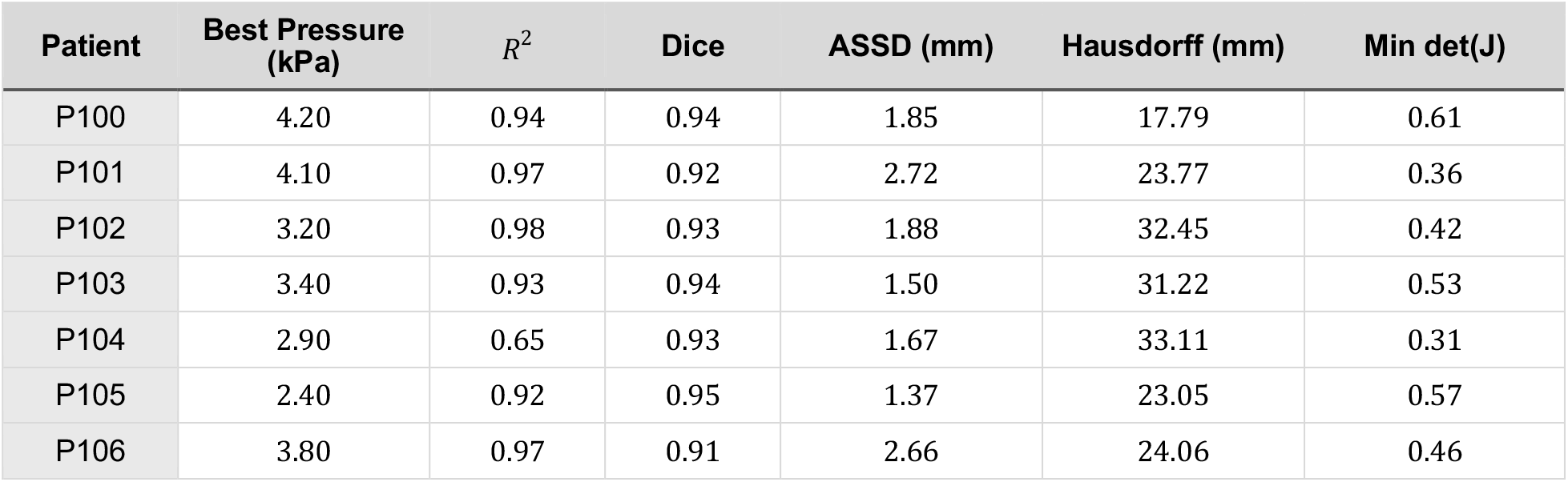
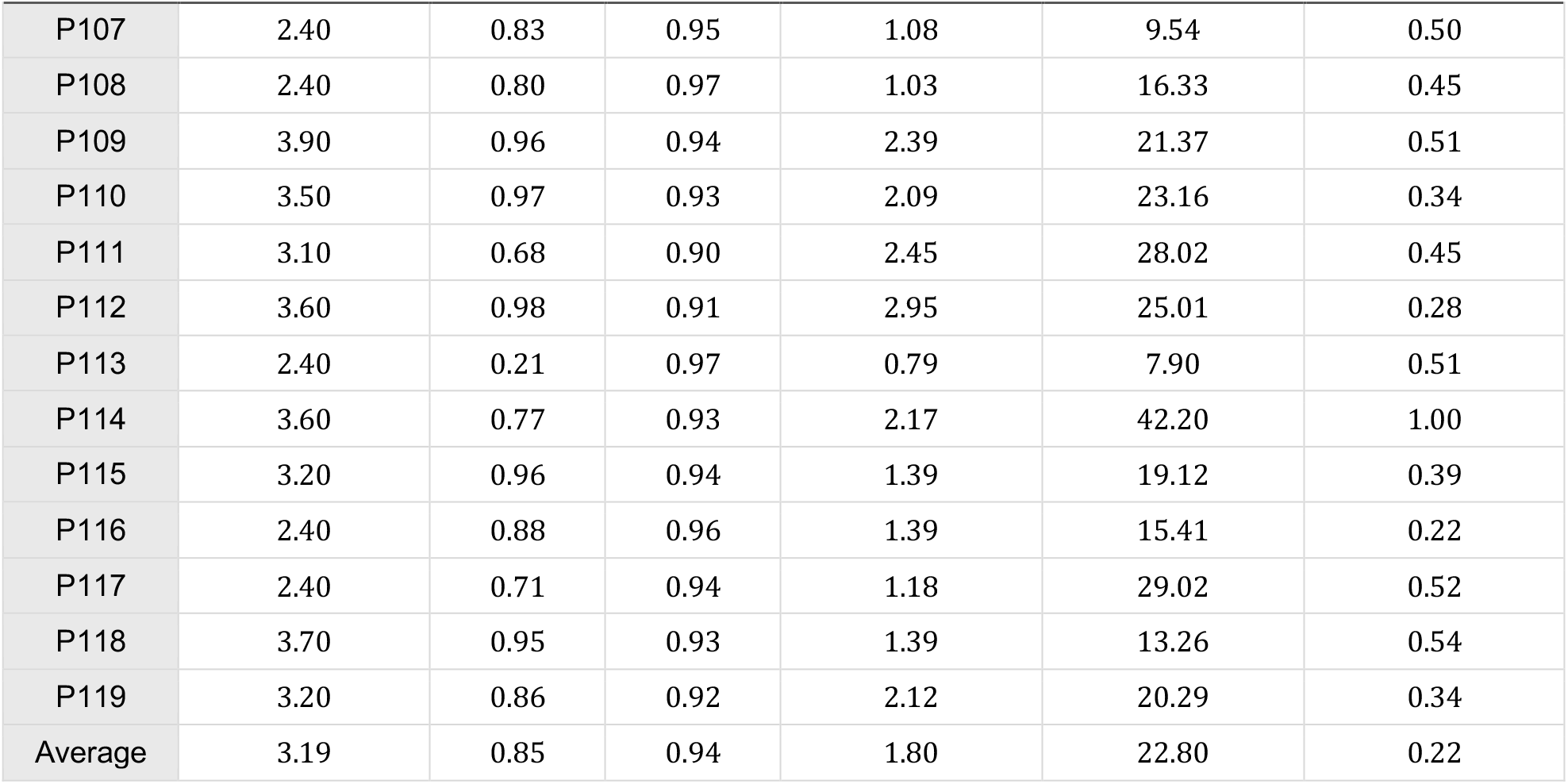
Measures of agreement between simulated and ground-truth lung masks across all 20 cases of the TCIA 4D-CT dataset. For each patient, we report the optimized intrathoracic pressure (kPa) used to match the lung-volume trajectory (with R^$^) and the worst-case mask agreement across respiratory phases (Dice, ASSD, and Hausdorff distance). The final row reports the mean across patients for all metrics except Min det(J), where it reports the minimum value.

| Patient | Best Pressure (kPa) | $R^2$ | Dice | ASSD (mm) | Hausdorff (mm) | Min det(J) |
| --- | --- | --- | --- | --- | --- | --- |
| P100 | 4.20 | 0.94 | 0.94 | 1.85 | 17.79 | 0.61 |
| P101 | 4.10 | 0.97 | 0.92 | 2.72 | 23.77 | 0.36 |
| P102 | 3.20 | 0.98 | 0.93 | 1.88 | 32.45 | 0.42 |
| P103 | 3.40 | 0.93 | 0.94 | 1.50 | 31.22 | 0.53 |
| P104 | 2.90 | 0.65 | 0.93 | 1.67 | 33.11 | 0.31 |
| P105 | 2.40 | 0.92 | 0.95 | 1.37 | 23.05 | 0.57 |
| P106 | 3.80 | 0.97 | 0.91 | 2.66 | 24.06 | 0.46 |
| P107 | 2.40 | 0.83 | 0.95 | 1.08 | 9.54 | 0.50 |
| P108 | 2.40 | 0.80 | 0.97 | 1.03 | 16.33 | 0.45 |
| P109 | 3.90 | 0.96 | 0.94 | 2.39 | 21.37 | 0.51 |
| P110 | 3.50 | 0.97 | 0.93 | 2.09 | 23.16 | 0.34 |
| P111 | 3.10 | 0.68 | 0.90 | 2.45 | 28.02 | 0.45 |
| P112 | 3.60 | 0.98 | 0.91 | 2.95 | 25.01 | 0.28 |
| P113 | 2.40 | 0.21 | 0.97 | 0.79 | 7.90 | 0.51 |
| P114 | 3.60 | 0.77 | 0.93 | 2.17 | 42.20 | 1.00 |
| P115 | 3.20 | 0.96 | 0.94 | 1.39 | 19.12 | 0.39 |
| P116 | 2.40 | 0.88 | 0.96 | 1.39 | 15.41 | 0.22 |
| P117 | 2.40 | 0.71 | 0.94 | 1.18 | 29.02 | 0.52 |
| P118 | 3.70 | 0.95 | 0.93 | 1.39 | 13.26 | 0.54 |
| P119 | 3.20 | 0.86 | 0.92 | 2.12 | 20.29 | 0.34 |
| Average | 3.19 | 0.85 | 0.94 | 1.80 | 22.80 | 0.22 |

**Table 9.** Ablation study: Measures of agreement between simulated and ground-truth lung masks across all 20 cases without pressure fitting (fixed to 2.4 kPa) and no global alignment. The final row reports the mean across patients for all metrics except Min det(J), where it reports the minimum value.

| Patient | Pressure (kPa) | Dice | ASSD (mm) | Hausdorff (mm) | Min det(J) |
| --- | --- | --- | --- | --- | --- |
| P100 | 2.40 | 0.94 | 1.87 | 17.34 | 0.77 |
| P101 | 2.40 | 0.96 | 1.22 | 16.42 | 0.50 |
| P102 | 2.40 | 0.94 | 1.64 | 25.57 | 0.53 |
| P103 | 2.40 | 0.95 | 1.46 | 31.77 | 0.66 |
| P104 | 2.40 | 0.94 | 1.67 | 32.95 | 0.37 |
| P105 | 2.40 | 0.95 | 1.37 | 23.05 | 0.57 |
| P106 | 2.40 | 0.93 | 1.36 | 16.33 | 0.56 |
| P107 | 2.40 | 0.95 | 1.08 | 9.54 | 0.50 |
| P108 | 2.40 | 0.97 | 1.03 | 16.33 | 0.45 |
| P109 | 2.40 | 0.89 | 2.82 | 36.79 | 0.66 |
| P110 | 2.40 | 0.93 | 2.14 | 22.60 | 0.49 |
| P111 | 2.40 | 0.91 | 2.29 | 28.34 | 0.52 |
| P112 | 2.40 | 0.95 | 1.34 | 21.93 | 0.39 |
| P113 | 2.40 | 0.97 | 0.79 | 7.90 | 0.51 |
| P114 | 2.40 | 0.94 | 2.03 | 42.44 | 0.42 |
| P115 | 2.40 | 0.94 | 1.36 | 19.49 | 0.31 |
| P116 | 2.40 | 0.96 | 1.39 | 15.41 | 0.22 |
| P117 | 2.40 | 0.94 | 1.18 | 29.02 | 0.52 |
| P118 | 2.40 | 0.93 | 1.41 | 13.07 | 0.64 |
| P119 | 2.40 | 0.92 | 2.09 | 20.05 | 0.45 |
| Average | 2.40 | 0.94 | 1.58 | 22.32 | 0.22 |

**Table 10.** Comparison of lung deformation accuracy across physics-based simulation and biomechanically regularized deformable registration methods evaluated on 4D CT. Ours is a forward finite-element breathing simulation validated on gated TCIA 4D-CT lung masks. Hering and Rühaak evaluate physics-regularized deformable lung CT registration on COPDGene inspiration/expiration scans. Jafari et al. present a forward hyperelastic finite-element lung tumor motion model evaluated on private clinical 4D-CT using tumor volumetric Dice. Min det(J) reports the minimum determinant of the deformation Jacobian across the evaluated TCIA cases for our best-pressure setting.

| Study | Year | Validation Dataset | Method | Dice | ASSD (mm) | HD (mm) | Min det(J) |
| --- | --- | --- | --- | --- | --- | --- | --- |
| Ours | 2026 | TCIA 4D-CT | Simulation | 0.94<br>$\pm 0.02$ | 1.80<br>$\pm 0.62$ | 22.80<br>$\pm 8.44$ | 0.22 |
| Hering et al. | 2021 | COPDGene | Registration | 0.95<br>$\pm 0.03$ | 1.72<br>$\pm 0.89$ | 26.8<br>$\pm 14.27$ | – |
| Rühaak et al. | 2017 | COPDGene | Registration | 0.92<br>$\pm 0.05$ | 1.97<br>$\pm 1.24$ | 27.24<br>$\pm 13.70$ | – |
| Jafari et al. | 2021 | Private clinical 4D-CT | Simulation | 0.85<br>$\pm 0.06$ | – | – | – |

## 4 Discussion

Across three modalities, mechanistic simulation produces imaging acquisitions rare or absent in standard clinical practice. Since generation follows from a constrained physical model rather than learned image synthesis, the method requires no training data, no paired examples, and no generative model, bringing patient-specific acquisitions within reach. The simulated respiratory cycle reproduced gated 4D CT closely, and the compression states were indistinguishable from real images to expert readers.

Quantitatively, physics-based augmentation consistently improved Dice scores relative to no augmentation and increased performance when combined with other transformations. The strongest performance was achieved when physics-based augmentation was paired with simple transformations such as flipping, outperforming other more complex deformation techniques such as CutMix or Elastic Deformation. Furthermore, these gains generalized across multiple backbones and architectures, indicating that improvements are not tied to a specific network family.

Physics-based synthetic data generation provides a practical mechanism to enrich datasets with controlled, mechanistically grounded variation that is difficult to acquire at scale in clinical practice. Computationally, the pipeline runs on standard CPU hardware, with a single simulation supplying an entire cycle at ∼20 s per generated image (Section 6.2.5). Varying simulation parameters within physiological bounds enables systematic exploration of rare anatomical configurations and acquisition conditions, selected deliberately rather than encountered by chance.

This approach may also be valuable for rare diseases, where few available cases limit the development of imaging AI. By simulating physiologic and acquisition variability around each observed case, mechanistic augmentation could expand the range of conditions under which a model encounters an uncommon pathology, extending usage of a scarce annotated data. Although these images do not replace independent patients or introduce new disease phenotypes, they may improve robustness to changes in positioning, compression, or motion that are poorly represented in small datasets.

Evaluating this potential in rare disease cohorts is an important direction for future work.

Several limitations warrant consideration. Physics-based synthetic data generation deforms existing anatomy rather than generating entirely novel structures, which may limit out-of-distribution generalization. The biomechanical models also rely on simplifying assumptions, such as rigid ribs, that introduce potential inaccuracies.

Material properties were taken from published values rather than measured per patient. Further, each modality was only evaluated on a single public dataset.

Overall, mechanistic simulation offers a route to imaging data that cannot otherwise be obtained, producing physiologically accurate imaging acquisitions extrapolated from standard imaging, with its effectiveness holding across modalities, architectures, and transformations.

## 5 Conclusion

We demonstrate that mechanistic simulation generates imaging acquisitions rare in standard clinical practice—a full respiratory cycle from a single breath-hold CT, a range of breast compressions from a single tomosynthesis volume, arterial wall distension across the cardiac cycle from a single carotid ultrasound frame — without training data, paired examples, or additional patient exposure. Where a reference standard exists, the generated motion matches real gated 4D CT; where none exists, expert readers cannot distinguish the result from real images. Training on these acquisitions improves lesion detection and segmentation across modalities, indicating that physiologically constrained synthesis supplies variation that neither standard transformations nor generative models provide.

## Supporting information

supplemental gifs

## Data Availability

This study used publicly available medical imaging data: the Breast Cancer Screening, Digital Breast Tomosynthesis (BCS-DBT) collection from The Cancer Imaging Archive (TCIA); the Lung-Originated Tumor Segmentation (LOTUS) benchmark, derived from NSCLC-Radiomics with additional annotations from the MAASTRO clinic; the Carotid Ultrasound Boundary Study (CUBS) dataset, accessed via Kaggle and described in the cited original dataset publication; and gated 4D chest CT data from TCIA for respiratory-motion validation. We also generated synthetic, physics-based augmented images and corresponding segmentation masks from these publicly available datasets by simulating breast compression, respiratory motion, and carotid arterial wall distension. The original datasets are available from their respective repositories under the applicable access requirements and terms of use. Key supporting code and data are provided in the Supplementary Information ZIP; the complete collection of generated synthetic data and code is not deposited in a public repository. Further methodological details are available from the corresponding author on request.

https://doi.org/10.7937/E4WT-CD02

https://i-sip.encs.concordia.ca/DataSets/LotusBenchmark/

https://doi.org/10.17632/m7ndn58sv6.1

https://www.cancerimagingarchive.net/collection/4d-lung/

## Author Contributions

C.S. developed the physics models, designed the study, and wrote the manuscript. A.D. performed lung preprocessing, prepared visualizations, and validated simulated lung motion against 4D CT. N.S.H. performed model ablation. A.S. segmented breast tumor structures. A.C. contributed to manuscript writing and segmentation corrections. K.W. performed statistical analysis of the reader study. I.L.X. downloaded and preprocessed the lung dataset and reviewed segmentations. M.V.P. developed code for segmentation pipeline. K.H. provided expert radiology guidance on manuscript writing and reviewed segmentations. P.S. contributed to breast segmentation generation and guidance on manuscript writing. M.H. (Marc Hirschvogel) generated and advised on arterial simulations. W.Z. revised and corrected segmentations. C.L. performed ablation studies. V.G. revised segmentations. A.G.E., M.S., W.V.C., and J.I.A. contributed to manuscript writing. T.N.N. contributed to physics modeling. L.S., A.C.I., F.I.-S., A.O.N., D.L.L., M.H. (Meng Hao), E.S.M., N.C.A., N.I.I., S.P.P., B.Z.D., S.M.D., and I.O. participated in the reader study. O.G. supported scientific direction.

### Funding

This research received no specific funding.

### Competing interests

The authors declare no competing interests.

### Data availability

This study used publicly available medical imaging data from the BCS-DBT collection (https://doi.org/10.7937/E4WT-CD02), the LOTUS benchmark (https://i-sip.encs.concordia.ca/DataSets/LotusBenchmark/), and the Carotid Ultrasound Boundary Study (CUBS; https://doi.org/10.17632/m7ndn58sv6.1), accessed via Kaggle as described in the Methods. Gated 4D chest CT data from TCIA were used for respiratory-motion validation.

Original data are available from their respective repositories under the applicable access requirements and terms of use. The Supplementary Information ZIP contains breast and lung simulation animations, a pressure-fitting animation, the reader-study quiz, and example lung augmentation code. The complete generated synthetic datasets and full code collection are not deposited in a public repository. Further methodological details are available from the corresponding author on request.

## 6 Supplemental Material

### 6.1 Extended Methods

#### 6.1.1 Datasets

#### 6.1.2 Tissue Properties

#### 6.1.3 Fluid-Structure Interaction (FSI)

Fluid-structure interaction (FSI) is used to describe the deformation of a vessel subject to (steady or pulsatile) blood flow. The vessel wall’s displacement ***u*** from its reference domain 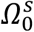 to its current, deformed configuration *Ω*^*s*^ is governed by the balance of linear momentum of finite strain elastodynamics,

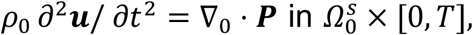

where *ρ*_0_ is the wall density and ***P*** the first Piola-Kirchhoff stress tensor. Its constitutive equation here is of hyperelastic type,

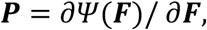

where ***F*** = ***I*** + ∇_0_***u*** is the deformation gradient and *ψ* the strain energy function. Here, an isotropic compressible Neo-Hookean model [15] is used.

The fluid is governed by the incompressible non-conservative Navier-Stokes equations in an Arbitrary Lagrangian-Eulerian (ALE) reference frame [10, 11], defining the conservation of linear momentum and mass over the current domain *Ω*^*f*^:

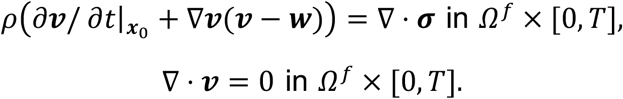

The fluid’s velocity and pressure are ***v*** and *p*, respectively. The fluid domain is displaced according to the exchange of forces between blood and tissue, characterized by the domain velocity ***w***. The domain motion is typically governed by a simple diffusion or pseudo-solid model [30]. The blood density is denoted by *ρ*. The blood is assumed to be a Newtonian fluid, making the Cauchy stress

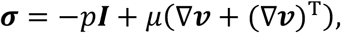

with the dynamic viscosity *µ*.

At the FSI interface *Σ* (mutual boundary shared by fluid and solid domain), both kinematic continuity and continuity of tractions have to hold [22, 24]:

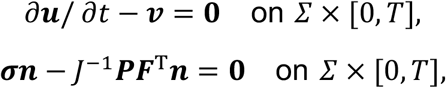

where ***n*** is a unit outward normal in the current frame (defined to point outward of the fluid domain) and *J* = det***F***.

The blood is further subject to a resistance boundary condition at the outflow boundary *Γ*^*R*^,

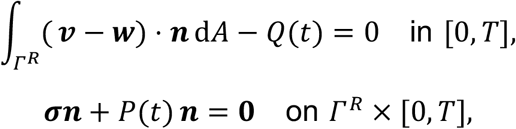

where the flow-pressure relationship of the resistor (parameter *R*) is given by Poiseuille’s law [33]:

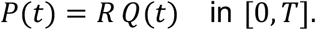

At the inflow boundary *Γ*^23^, the velocity is controlled by a parabolic Dirichlet condition with mean value ***v*** *⋅* ***n*** = 85 cm/s.

Solid and fluid are discretized in space with P_2_-P_1_Taylor-Hood finite elements [34] with the resulting mesh shown in Figure 1I. Time discretization is performed using the Generalized-*α* method [9, 17].

The resulting discretized system of equations is solved with the open-source multi-physics solver Ambit, licensed under LGPL v3 [14], which uses the finite element backend FEniCSx version 0.7.3, licensed under LGPL v3 [3, 19] and linear algebra suite PETSc licensed under the BSD 2-Clause license [2].

#### 6.1.4 Lung Validation

##### 6.1.4.1 Dataset

The 4D-CT dataset comprises 20 patients with tumors, with scans acquired on multiple dates at 10 distinct phases of the respiratory cycle. The 4D-CT enables direct comparison of simulated respiratory deformation with ground-truth motion at each of the 10 phases. All 20 cases were used for validation.

##### 6.1.4.2 Segmentation

Lung and bone structures were segmented using TotalSegmentator, following the same procedure described in Section 2.2.1. Tumor segmentations were obtained by applying the MONAI U-Net model, which we finetuned for predicting gross tumor volumes in the LOTUS dataset [8]. Similarly, the body segmentation was generated by implementing a separate MONAI U-Net model, trained on the LOTUS dataset’s body segmentations. To avoid inclusion of irrelevant background regions, a bounding box encompassing all relevant organs was constructed using TotalSegmentator outputs. The predicted body segmentation was then cropped to this bounding region.

All 20 patients’ segmentations (lung, bone, tumor, and body) were reviewed by a clinician to verify anatomical correctness and suitability for simulation.

##### 6.1.4.3 Fitting

To account for patient-specific respiratory parameters, we performed a grid search over intrathoracic pressure values ranging from 2400 Pa to 4500 Pa. Simulated lung configurations were compared against the corresponding ground-truth gated respiratory phases. Prior to evaluation, a translational alignment was applied to maximize spatial overlap between the simulated and ground-truth lung volumes, compensating for global patient motion (e.g., table or body shifts) not modeled by the simulation. Pressure values were selected by maximizing the coefficient of determination (*R*^2^) between simulated and ground-truth lung volume curves over the respiratory cycle, with the best-fitting value used for subsequent metric evaluation.

### 6.2 Extended Results

#### 6.2.1 Qualitative Comparison of augmentation strategies

#### 6.2.2 Reader Study: Simulation Artifacts identified by reader 3

Rounding of sharp tissue boundaries at the image edges during mesh generation can introduce minor border artifacts, which can be mitigated by enforcing planar constraints during meshing, as supported by existing functionality in meshing software. Skin markers, which consist of a thin metallic ring, were not modeled explicitly and therefore deform according to surrounding soft-tissue parameters, leading to unnatural elongation and blurring of their usual delineation. Here incorporating marker-specific material properties would address this.

#### 6.2.3 Lung Validation

#### 6.2.4 p-value multiple hypothesis correction

In order to view the effect of physics-based augmentation, we compared physics-based augmentation training against their matched non-physics counterparts within the same augmentation setting. For each matched comparison, we used a two-sided Welch’s t-test which were applied to their respective dice metrics. Since we conduct multiple hypothesis tests for the dice metric, we controlled for multiple comparisons using the Benjamini-Hochberg false discovery rate (FDR) procedure. FDR correction was applied only on a per dataset and per metric basis, so the adjustment for tumor dice in the breast dataset depends only on the set of tumor-dice comparisons within that dataset and vice versa for the other datasets.

#### 6.2.5 Run time analysis

For the lung dataset, the full generation pipeline was executed on CPU (AMD Threadripper 7965WX, 24 cores). Per case, segmentation took ∼1 min using TotalSegmentator, mesh generation ∼2 min, simulation ∼2 min, and applying the deformation field ∼3 min, totaling ∼8 min per 3D volume. For 292 cases, this corresponds to approximately 1.5 days of wall-clock time without specialized hardware such as GPUs. Critically, a single simulation produces multiple augmented samples (e.g., 24 respiratory time steps per breathing cycle), amortizing the per-sample cost and enabling diverse augmentations without repeated simulation.

## Notes

### Competing Interest Statement

The authors have declared no competing interest.

### Author Declarations

The study used publicly available human imaging datasets released before study initiation: BCS-DBT (https://doi.org/10.7937/E4WT-CD02), LOTUS (https://i-sip.encs.concordia.ca/DataSets/LotusBenchmark/), CUBS (https://doi.org/10.17632/m7ndn58sv6.1; accessed via Kaggle as described in the manuscript), and TCIA 4D-Lung (https://www.cancerimagingarchive.net/collection/4d-lung/). Biomechanical simulations generated additional images and segmentation masks from these existing datasets. No new patient imaging or prospective treatment assignment was performed.

