## Supplementary figures and images for "Training-free generation of 4D CT and other rare acquisitions improves segmentation across modalities"

### breast.gif

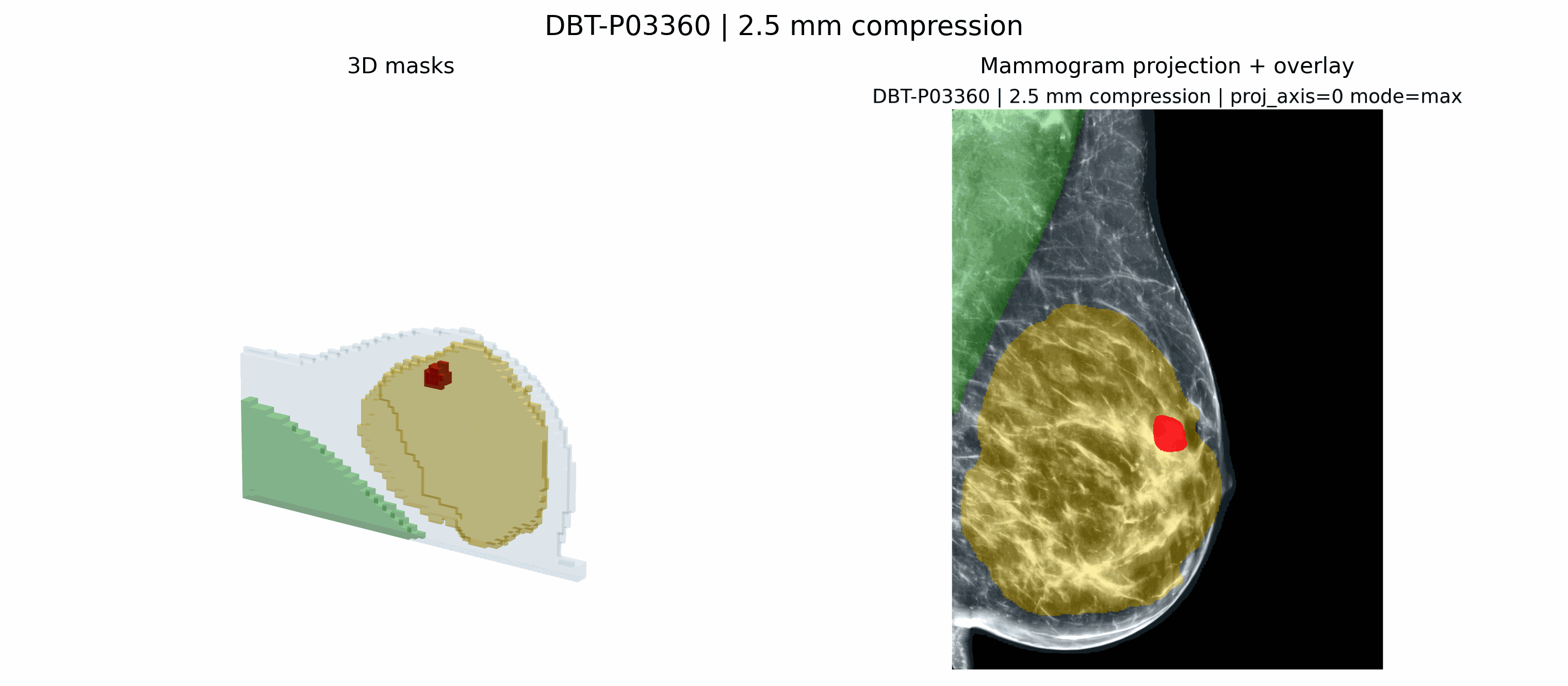

### lung.gif

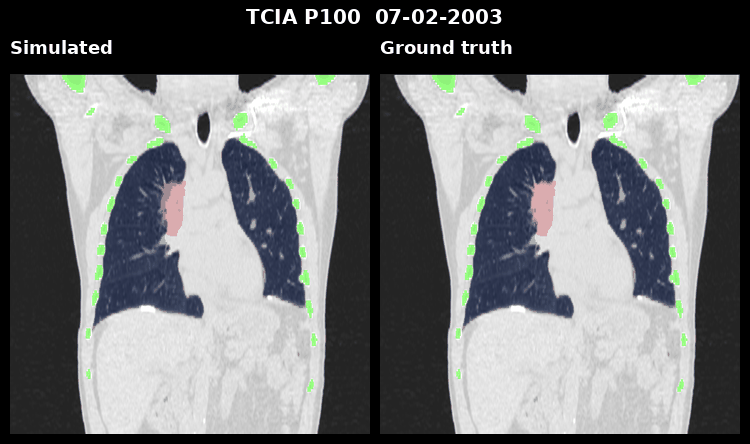

### pressure_fitting.gif

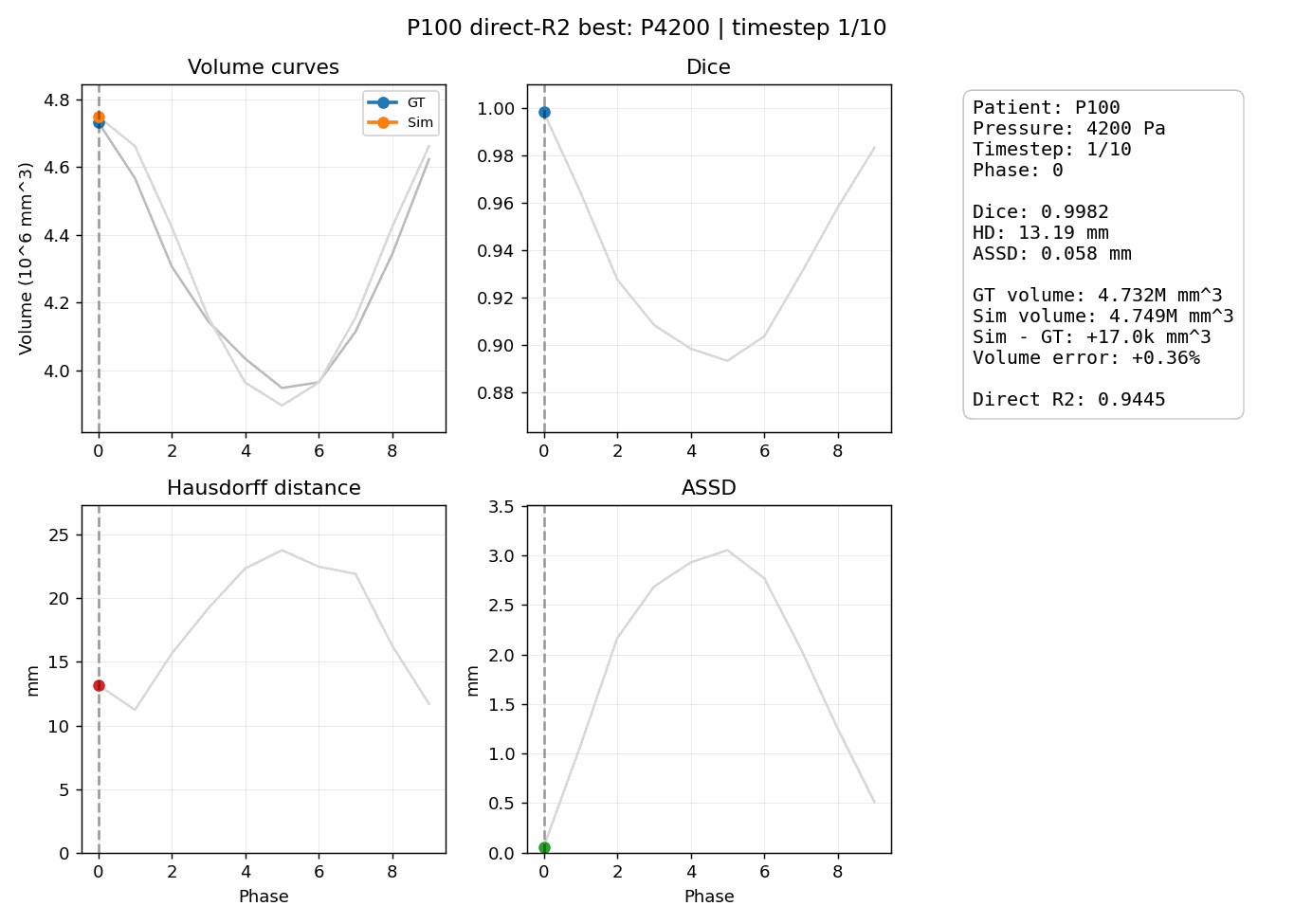
